# Street images classification according to COVID-19 risk in Lima, Peru: A convolutional neural networks analysis

**DOI:** 10.1101/2021.09.06.21263188

**Authors:** Rodrigo M Carrillo-Larco, Jose Francisco Hernández Santa Cruz

## Abstract

**Background:** During the COVID-19 pandemic, convolutional neural networks (CNNs) have been used in clinical medicine (e.g., to classify chest X-rays for COVID-19 diagnosis). Whether CNNs could also inform the epidemiology of COVID-19 analysing street images has been understudied, though it could identify high-risk places and relevant features of the built environment. We trained CNNs to classify bus stops (Lima, Peru) into moderate or extreme COVID-19 risk.

**Methods:** We used five images per bus stop. The outcome label (moderate or extreme) for each bus stop was extracted from the local transport authority. We used transfer learning and updated the output layer of five CNNs: NASNetLarge, InceptionResNetV2, Xception, ResNet152V2, and ResNet101V2. We chose the best performing network which was further tuned to increase performance.

**Results:** There were 1,788 bus stops (1,173 moderate and 615 extreme), totalling 8,940 images. NASNetLarge outperformed the other CNNs except in the recall metric for the extreme label: 57% versus 59% in NASNetLarge and ResNet152V2, respectively. NASNetLarge was further tuned and reached: training loss of 0.50; training accuracy of 75%; precision, recall and F1 score for the moderate label of 80%, 83% and 82%, respectively; these metrics for the extreme label were 65%, 51% and 63%.

**Conclusions:** CNNs has the potential to accurately classify street images into levels of COVID-19 risk. In addition to applications in clinical medicine, CNNs and street images could also advance the epidemiology of COVID-19 at the population level.

## INTRODUCTION

In COVID-19 research, deep learning tools applied to image analysis (i.e., computer vision) have informed the diagnosis and prognosis of patients through classification of chest X-ray and computer tomography images.^1-3^ These tools have greatly helped practitioners treating COVID-19 patients.

On the other hand, the application of computer vision to study the epidemiology of COVID-19 has been limited. One relevant example is the use of Google Street View images to extract features of the built environment and associate these with COVID-19 cases in the United States.^4^ This work showed that unstructured and non-conventional data sources, such as street images, can deliver relevant information to characterize the epidemiology of COVID-19 at the population level.^4^ However, computer vision models to classify street images based on their COVID-19 risk do not exist. These models could be relevant to understand unique local features of the built environment related to high COVID-19 risk. In addition, these models could be applied to places where observed data are not available to identify whether this place is at moderate or high risk of COVID-19, and then receive interventions as needed. This could be particularly helpful in Low- and Middle-income countries where limited resources do not allow massive COVID-19 testing, leaving places with no information about the COVID-19 epidemiology.

In this feasibility study, we aimed to ascertain whether a convolutional neural network (deep learning) model could correctly classify street images of bus stops according to their COVID-19 risk (binary outcome: moderate versus extreme risk) in Lima, Peru. We also aimed to understand what features of the images were most influential in the classification process.

## METHODS

### Study design

We used convolutional neural networks to study street images of bus stops in Lima, Peru. We implemented a classification model to categorize the bus stops into two labels: moderate or extreme risk of COVID-19.

### Data sources

The labels (observed data) of the bus stops were downloaded from the website of the Authority for Urban Transport in Lima and Callao (*Autoridad de Transporte Urbano para Lima y Callao* (ATU), name in Spanish). This government office manages the public transportation service in Lima, and publishes a classification map in which all bus stops in Lima are set into four categories of COVID-19 risk: moderate < high < very high < extreme.^5^ Further details of their classification system have not been released. In this feasibility study, we only worked with the bus stops deemed as moderate (label 0) and extreme (label 1) risk of COVID-19. We used the classification profile released on 2021-05-24^.6^

We used the location (longitude and latitude coordinates) of the bus stops to download their street images through the application programming interface (API) of Google Street View. For each bus stop (i.e., from each coordinate), we downloaded five images: when the camera was facing at 0 degrees, at 90 degrees, at 180 degrees and at 270 degrees; in addition, we also downloaded one image in which the direction of the camera was not specified (i.e., the heading parameter in the API request was set at default). In other words, for each bus stop we had five images. We did this to maximise the available data and to cover the surrounding area of the bus stop.^7^ Our rationale was that the bus stop itself would not be responsible for the classification, but the whole nearby environment.

We combined the images and the labels in one dataset, which was further divided into three datasets: the training dataset including 60% of the data, the validation dataset including 20%, and the test dataset including the remaining 20%. The allocation to each of these three datasets was at random.

### Analysis

In-depth details about the analysis are available in Supplementary Materials pp. 03-06. The analysis code (Python Jupyter notebooks) is also available as supplementary materials.

In brief, in a pre-specified analysis we decided to elaborate on five deep convolutional neural networks pre-trained with ImageNet (i.e., transfer learning). We chose these models because they have the best top-5 accuracy of the models available in the Keras library:^8^ NASNetLarge, InceptionResNetV2, Xception, ResNet152V2 and ResNet101V2. We implemented these five models with the same hyper-parameters, and then we selected the one with the best performance which was further tuned and tested. The image classification model was based on the latter model only (i.e., the one with the best performance out of the five candidate models). We reported the loss and accuracy in the validation and test datasets; we also used the test dataset to report the accuracy, recall and F1 score for each of the two possible outcomes (moderate or extreme risk). Finally, we used GradCam to identify which areas of the input image were more relevant to inform the classification;^9^ for this, we randomly selected four images per outcome (i.e., four images from the moderate label and four images from the extreme label).

### Ethical statement

Human subjects were not directly studied in this work. The analysed data are in the public domain and can be downloaded. The analysed data do not contain any personal identifiers. We did not seek approval by an ethics committee.

## RESULTS

### Available data

There were 1,788 bus stops with their corresponding label: 1,173 in the *moderate* category and 615 in the *extreme* category. Because we used five images per bus stop, the analysis included 8,940 (1,788×5) images and their corresponding label. The training dataset included a random sample of 60% (5,364) of the original dataset, while the validation and test datasets included a random sample of 20% (1,788) each.

### Selection of the pre-trained model out of five candidate models

We used transfer learning and updated the output layer of five convolutional neural networks to predict our two classes of interest. The NASNetLarge architecture and weights outperformed the other candidate convolutional neural networks, except in the recall metric for the *extreme* label: 57% versus 59% in NASNetLarge and ResNet152V2, respectively (Table 1). Further experiments were only conducted with NASNetLarge.

**Table 1:** Performance of the five candidate convolutional neural networks.

|  | NASNetLarge | InceptionResNetV2 | Xception | ResNet152V2 | ResNet101V2 |
| --- | --- | --- | --- | --- | --- |
| **Loss, validation** | 0.506450 | 0.533010 | 0.508640 | 0.612390 | 0.782688 |
| **Accuracy, validation** | 0.766477 | 0.737500 | 0.758522 | 0.746590 | 0.731818 |
| **Loss, test** | 0.509153 | 0.544169 | 0.524600 | 0.619583 | 0.777452 |
| **Accuracy, test** | 0.747727 | 0.728409 | 0.723863 | 0.733522 | 0.716477 |
| **Precision, label 0 (*moderate*)** | 0.79 | 0.78 | 0.78 | 0.79 | 0.76 |
| **Recall, label 0 (*moderate*)** | 0.84 | 0.81 | 0.81 | 0.81 | 0.83 |
| **F1 score, label 0 (*moderate*)** | 0.81 | 0.80 | 0.79 | 0.80 | 0.79 |
| **Precision, label 1 (*extreme*)** | 0.65 | 0.62 | 0.61 | 0.62 | 0.61 |
| **Recall, label 1 (*extreme*)** | 0.57 | 0.57 | 0.56 | 0.59 | 0.50 |
| **F1 score, label 1 (*extreme*)** | 0.61 | 0.59 | 0.58 | 0.60 | 0.55 |

Green colour highlights the best metric, yellow colour highlights the second best metric and red colour highlights the third best metric row-wise. The precision, recall and F1 score are presented as proportions (multiply by 100 to have percentages). The precision, recall and F1 score were computed with the test dataset.

### Model performance

We further tuned NASNetLarge with different hyper-parameters aiming to improve the accuracy (Table 2).

**Table 2:**
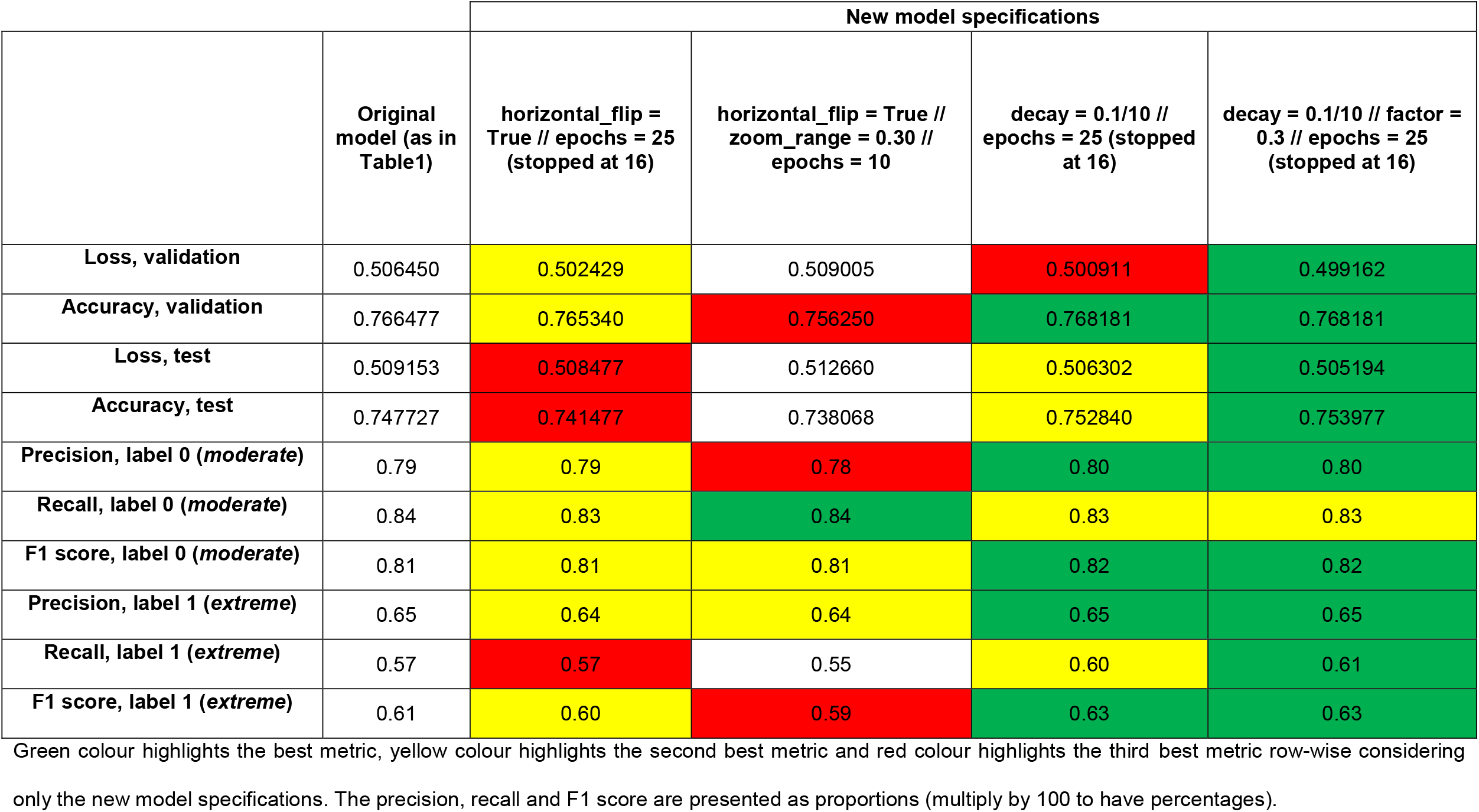
Further tuning of the selected model (NASNetLarge) and the performance metrics.

First, building on the initial hyper-parameters, we implemented two data augmentation options: horizontal flip and zoom range. We chose these two data augmentation methods because they appeared to fit the images under analysis; for example, because we were working with street images, a vertical flip would not seem appropriate. These data augmentation methods did not substantially improve the model accuracy.

Second, also building on the initial hyper-parameters (i.e., without data augmentation), the decay in the stochastic gradient descendent optimizer was changed from 1/25 (25 was the number of epochs) to 1/10 (the number of epochs was not changed). In this case, the model performance improved in comparison to the original model, particularly the metrics for the classification of the *extreme* label. For example, the recall metric for the *extreme* label was 57% in the original model and 60% with this new decay specification.

Third, building on the last specification (i.e., model with a decay of 1/10), we updated he monitoring factor which updated the learning rate when it did not improve through epochs. Originally, this factor was 0.1, and we updated it to 0.3. This new model had a marginal improvement versus the second one. For example, the recall metric for the *extreme* label was 57% in the original model, 60% in the second model (previous paragraph) and 61% in this third model (Figure 1). This last model was chosen, and implemented for the GradCam analysis.

**Figure 1:**
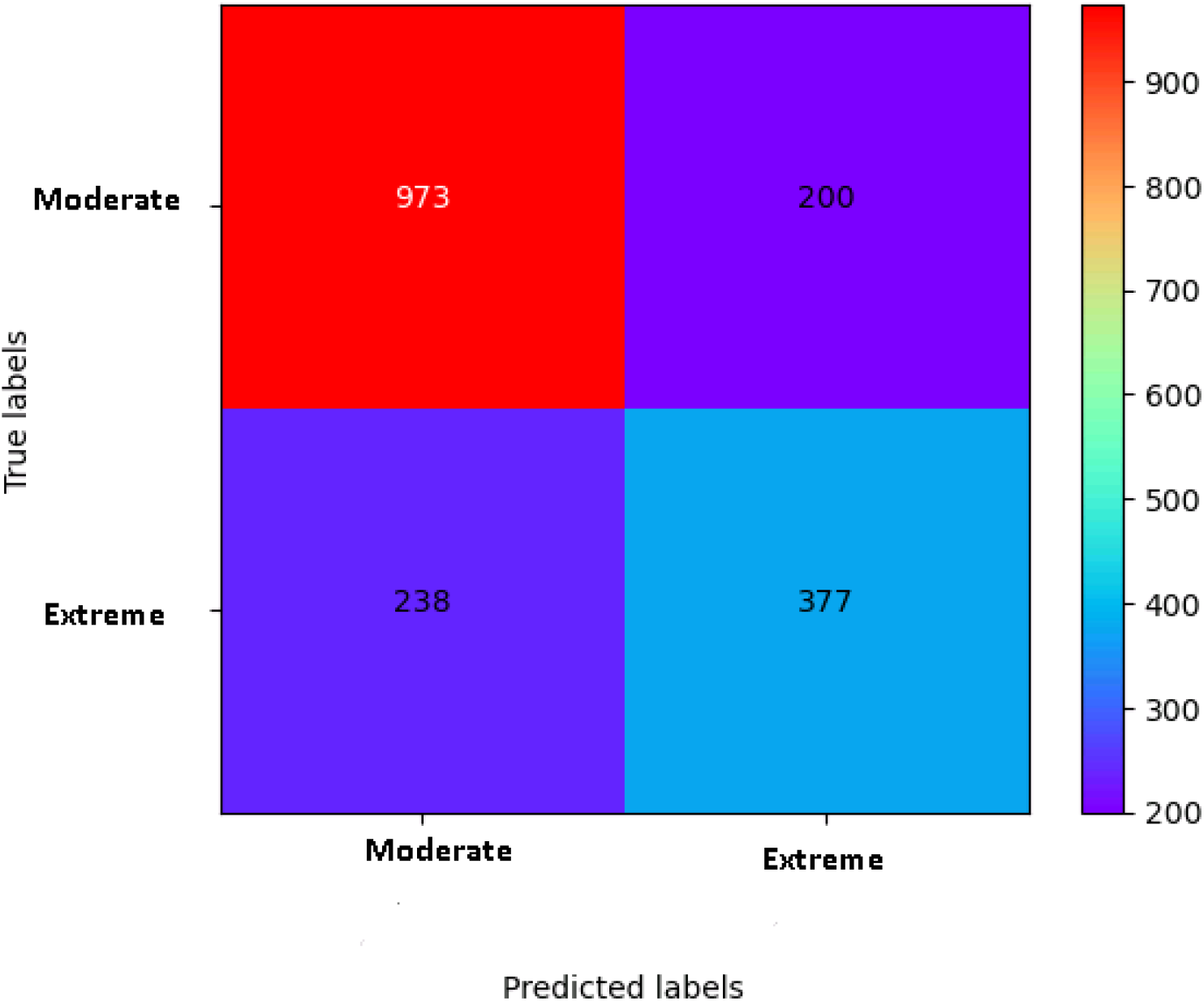
Confusion matrix for the best NASNetLarge model. This NASNetLarge model corresponds to the last column in Table 2. The figure shows the absolute number of images in each label: observed (true) on the y-axis and predicted on the x-axis.

### GradCam

The main indications for a moderate risk classification were the presence of green areas and lack of close nearby buildings (Figure 2). Areas close to buildings or with a considerable presence of people were often classified as extreme COVID-19 risk. The presence of cars did not seem to impact the classification process.

**Figure 2:**
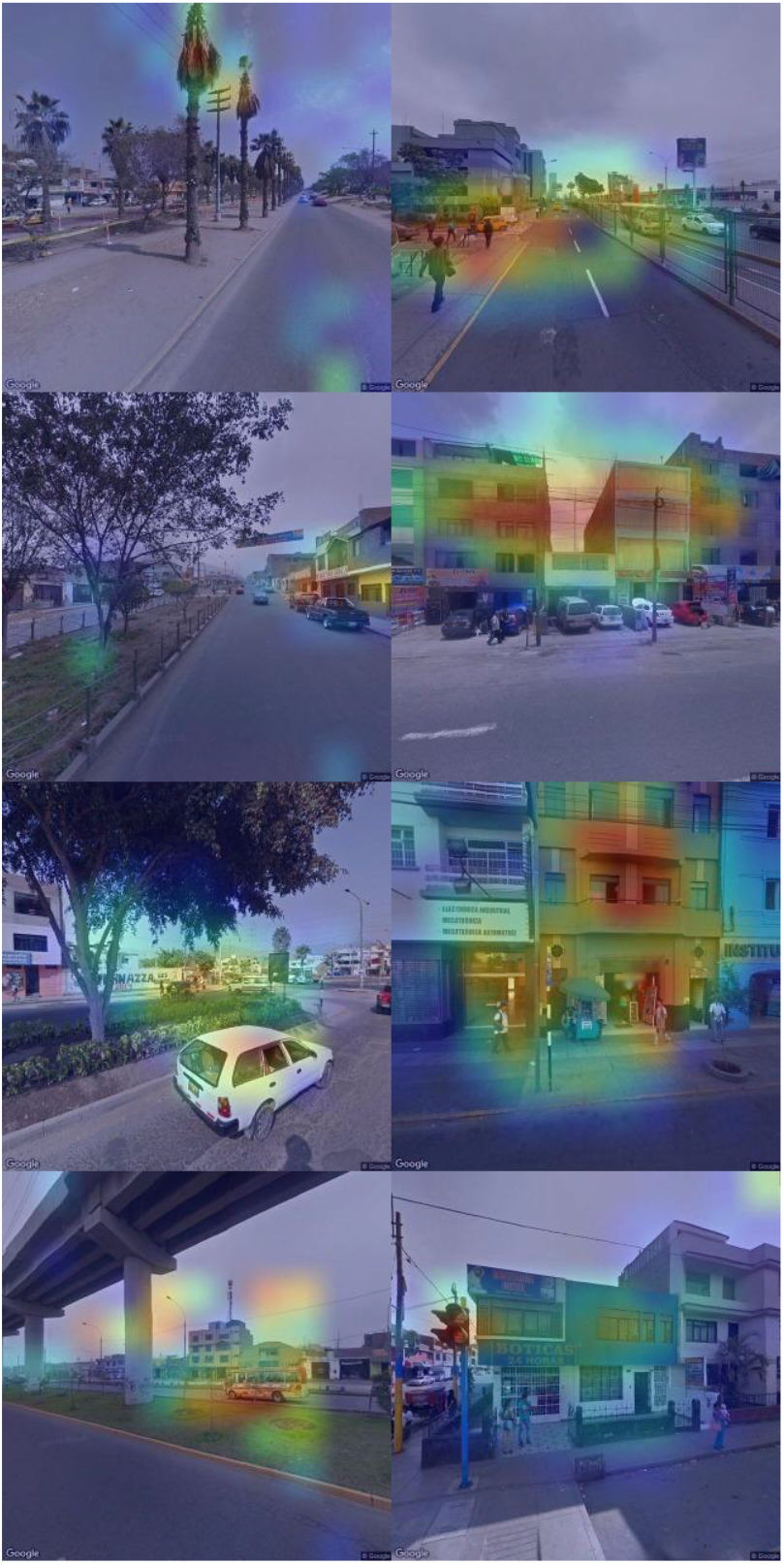
GradCam representation of four randomly selected images per outcome label (moderate risk on the left and extreme risk on the right) and based on the final model. Final model correspond to the NASNetLarge showed on the last column of Table 2. The four images per outcome label were randomly chosen. From green to red, it shows the areas that influenced the most in the classification process; that is, areas in red were the most influential in the classification process. Images on the left column belong to the moderate label and images on the right column belong to the extreme label. Images courtesy of Google Maps View.

## DISCUSSION

### Main findings

In this feasibility study, we showed that deep convolutional neural networks can classify street images according to their COVID-19 risk with acceptable accuracy. Future work should strengthen available convolutional neural networks or develop a new architecture which could maximize the accuracy classification, not only for a binary outcomes but also covering multiple outcomes. This work could spark interest to use convolutional neural networks –and other artificial intelligence tools– to advance population health and the epidemiological knowledge of COVID-19 (and other diseases), above and beyond the applications of convolutional neural networks for diagnosis and prognosis of individual patients (e.g., classification of chest X-rays and compute tomography images^1-3^).

### Results in context

This feasibility work signalled that a deep neural network is moderately accurate to classify street images according to COVID-19 risk levels. These results are encouraging because the task we pursued was difficult: to classify street images into levels for which there is no intrinsic information in the images. Classification of, for example, chest X-ray images into healthy or ill could be easier for a convolutional neural network because the X-ray of someone with a disease (e.g., pneumonia) would have unique features (e.g., infiltrate spots at the bottom of the lungs) that a healthy chest X-ray would not have at all. Conversely, in our case, the street images did not have a unique underlying pattern to guide the classification process; our model had to work harder to find those unique characteristics to decide between *moderate* and *extreme* risk.

Further tuning of the selected model (NASNetLarge) suggested that data augmentation methods did not improve the performance of the model; at least not the data augmentation options we implemented. This could suggest that for this particular task we may not need a very large number of images, but a careful consideration of the learning process in the network development and training. As a matter of fact, when we updated the learning rate optimizer, the model performance improved. Overall, these observations may imply that future work does not need to have many more images, but try different learning rates profiles or optimizers.

Nguyen and colleagues used Google Street View images to associate features of the built environment with COVID-19 cases in several states in the United States.^4^ Although we could have followed the same approach, there would be some unique local features of the built environment that may not have been identified by available object detection tools (e.g., street vendors and newspaper stands). We are not aware of other peer-reviewed papers in which street images have been classified in relation to COVID-19 outcomes. Our work contributes to the available literature with a newly trained model benefiting from transfer learning from a large and well-known architecture (NASNetLarge), based on images from a city in an upper-middle income country (Lima, Peru).

The activation maps (GradCam) are not only useful to analyse the model’s interpretation capability, but they bolster the existing evidence of crowded places or indoor venues (such as nearby buildings) as COVID-19 high risk areas; on the other hand, open spaces, such as green areas or locations far from buildings, remain as moderate or low COVID-19 risk areas. Overall, our findings agree with the evidence describing crowded areas, such as restaurants, gyms, hotels, and cafes, as having high COVID-19 infection risk^.10^

### Public health implications

Our work could have pragmatic applications to better understand the epidemiology of COVID-19 and to inform public health measures. For example, our model –and future work improving this analysis– could be used to characterize bus stops and other public places for which labelled data are not available. We worked with images from bus stops in Lima, and our model could be applied to bus stops in other cities to characterize their COVID-19 risk. Furthermore, our work could spark interest to conduct more sophisticated analyses, like semantic segmentation whereby some unique elements of the local environment could be identified as potential high-risk places. For example, bus stops often host food street vendors and newspaper stands where people usually gather. Perhaps, the bus stops themselves are not high-risk places, but the surrounding shops. This could inform policies and interventions to reduce the COVID-19 risk in these places. Overall, deep learning techniques, including convolutional neural networks, could be adopted by epidemiological research to advance the evidence about risk factors as well as disease outcomes and distribution, in addition to their current use in clinical medicine.^1-3^

### Ongoing and future work

The results showed a good accuracy to classify into the *moderate* label; probably because there were more bust stops under this classification. Future work should take under consideration the potential caveats of imbalanced data to improve the classification accuracy for the *extreme* risk label and other labels with fewer observations.

Ongoing and future work includes the development of a classification model for the four outcome labels (i.e., *moderate, high, very high* and *extreme* COVID-19 risk). We will implement techniques that can potentiate the classification capacity of the neural networks, including ensemble models,^11^ novel loss functions not currently implemented in the Keras environment (e.g., squared earth mover’s distance-based loss function),^12^ and we may try other architectures (e.g., SqueezeNet^13^) with similar precision yet less computationally expensive.

### Strengths and limitations

In this preliminary work, we followed a pre-defined analytical protocol which included transfer learning leveraging on large and deep neural networks trained with millions of images (ImageNet). We still had to train the parameters of the output layer, for which we did not have a massive number of images. Future work could expand our analysis with information and images from more bus stops or other public spaces to train a more robust model. Ideally, these images should come from different cities. This information may be available in other countries. There are further limitations we must acknowledge. First, the images and labels were not synchronic; that is, the figures and the labels were not collected on the same date. This is a shared limitation with other studies working with street images from open sources (e.g., Google Street View), because these images are not taken continuously or in real time. This should not be a major limitation because the analysis mostly focused on the built environment which has not changed substantially in recent years. Because this feasibility study showed that the classification model performed moderately well, researchers could collect new images in a prospective work to verify our findings with synchronic data. In this line, satellite images collected daily could be useful. Second, we did not have exact details on how the bus stops were classified by the local authorities. Nevertheless, we used official information which is provided to the public for their safety and to inform them about the progression of the COVID-19 pandemic in Lima, Peru. Therefore, we trust their method for classification is sound and based on the best available evidence. Third, we had five images per bus stop: the fifth image did not look at a specific angle, unlike the other four images that looked at 0, 90, 180 and 270 degrees around the bus stop. Therefore, the fifth image had some overlap with the other images. We took this decision to maximize the available data. Researchers with access to more labelled information, perhaps from public places overseas, could use the four images without overlap and not significantly reducing the dataset size.

## Conclusions

This feasibility study showed that a convolutional neural network has moderate accuracy to classify street images into *moderate* and *extreme* risk of COVID-19. In addition to applications in clinical medicine, deep convolutional neural networks have the potential to also advance the epidemiology of COVID-19 at the population level exploding unstructured and non-conventional data sources.

## Data Availability

All data sources and access means are described in the manuscript.

## CONTRIBUTIONS

RMC-L and JFHSC conceived the idea. RMC-L conducted the analysis with support from JFHSC. Both authors approved the submitted version.

## FUNDING

Wellcome Trust (214185/Z/18/Z)

## COMPETING INTERESTS

No conflict of interest.

## DATA AVAILABILITY STATEMENT

Outcome (i.e., labels: moderate and extreme COVID-19 risk) data are available online: https://sistemas.atu.gob.pe/paraderosCOVID; this information was systematized at https://github.com/jmcastagnetto/lima-atu-covid19-paraderos. The images were downloaded from Google Street View through the API with a personal account; images cannot be shared with third parties.. All analysis codes are available as Python Jupyter Notebooks in supplementary materials.

## Supplementary Materials

## EXPANDED METHODS

### Overview

In a pre-specified protocol we decided to test five well-known convolutional neural network architectures. These five networks are those with the best accuracy among the models available in the Keras library.^1^ From these five candidate models, we chose the one with the best performance for our task; this model was further tuned to improve its performance.

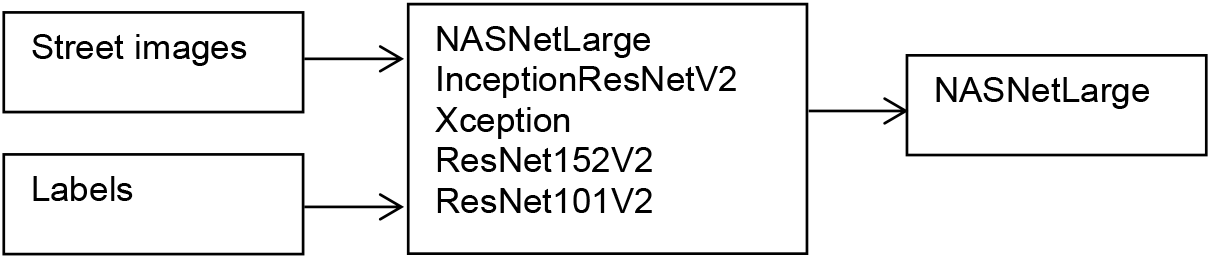

Analysis code (Jupyter notebooks) and the weights of the final model are found here: https://drive.google.com/file/d/1HXLsenn7yvxri7n2xE80WMtxQzj5fgBX/view?usp=sharing

### System details

Analyses were conducted with a GPU NVIDIA Quadro P1000 on Python Jupyter (version 3.7.10). The notebooks are provided as supplementary materials.

### Images

The list of bus stop in Lima, Peru, and their COVID-19 risk, are provided by local transport authority.^2^ This information has been extracted and homogenized and is available online.^3^ Key for our work, this information contains: i) location of each bus stop (latitude and longitude coordinates), which was used to extract street images; and ii) the COVID-19 risk level assigned to each bus stop, which was used as the outcome labels.

We downloaded the images from Google Street View through the application programming interface (API). We used the Python libraries *google_streetview*.*api* and *request*. For each bus stop (i.e., for each latitude and longitude coordinate), the API request specified the heading parameter = [0, 90, 180, 270]; in addition, we downloaded one image for which the heading parameter was set at default. Consequently, for each bus stop we had five images in total. The images were downloaded with size 640×640 pixels. In this feasibility study there were 1,788 bus stops, and because we had five images per bus stop, we used 8,940 (1,788 × 5) images in total.

### Labels (outcome)

We used the bus stop classification released on 2021-05-24,^2, 3^ by the local transport authority in Lima, Peru.^2^ They classify the bus stops in four categories of COVID-19 risk: moderate, high, very high and extreme risk.^2^ Details on how they made this classification are not available. Nevertheless, because this is official information released by a public authority to inform the general population, we trust their classification is based on the best available evidence. In this feasibility work we only used bus stops labelled as *moderate* (n=1,173) and *extreme* (n=615) COVID-19 risk. There were 1,788 (1,173 + 615) bus stops in total. The list of labels was appended five times, so that we would have as many images as labels (NB: we had five images per bus stop as described above).

### Image data generator

We constructed a dataframe with two columns: the path to each image (i.e., to the exact location were the images were saved) and the corresponding label for each image. This dataframe was passed to the *ImageDataGenerator* function of the *keras*.*preprocessing*.*function* library. At this point, we also re-scaled the images between 0 and 1 by dividing by 255. Then, we created three image iterators: one for the training, validation and test datasets (*train_datagen*.*flow_from_dataframe* function). To the *train_datagen*.*flow_from_dataframe* function we passed the datarame with the location of the images and their labels, specified this was a categorical classification problem, a batch size of 32, and a specific image size for each candidate model (see table below); in addition, the image iterator for the training dataset had the shuffle parameter as *True*.

### Convolutional neural networks

We decided to train five candidate convolutional neural networks with the following specifications.

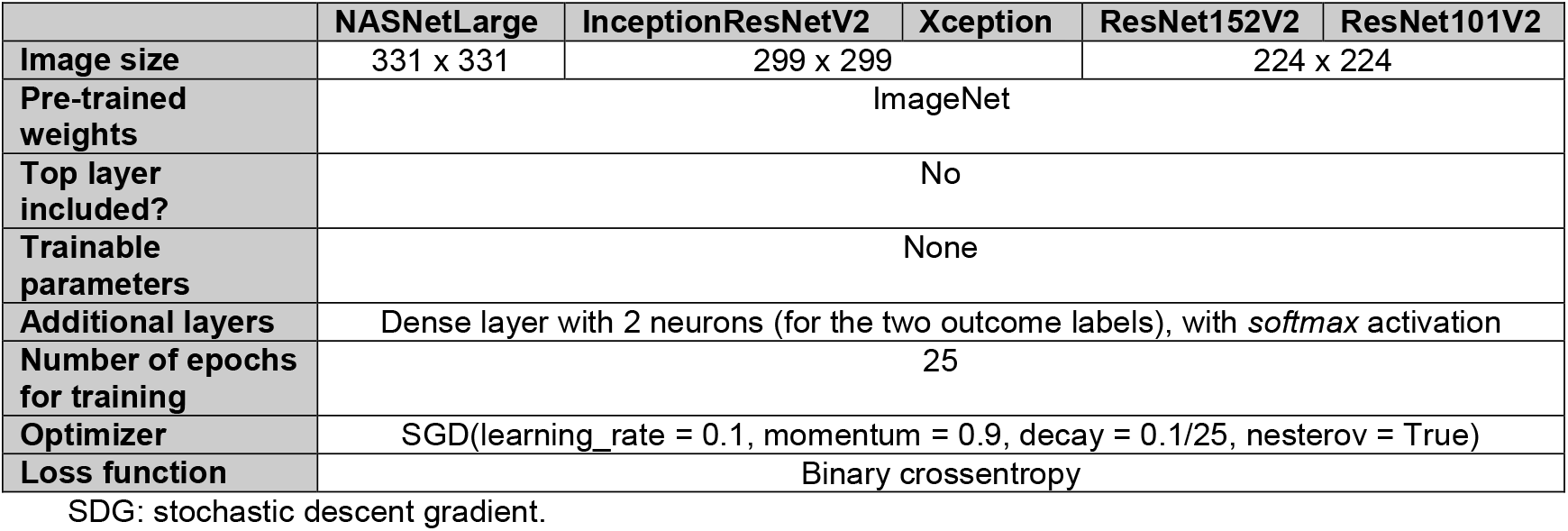

|  | NASNetLarge | InceptionResNetV2 | Xception | ResNet152V2 | ResNet101V2 |
| --- | --- | --- | --- | --- | --- |
| Image size | 331 x 331 | 299 x 299 |  | 224 x 224 |  |
| Pre-trained weights | ImageNet |  |  |  |  |
| Top layer included? | No |  |  |  |  |
| Trainable parameters | None |  |  |  |  |
| Additional layers | Dense layer with 2 neurons (for the two outcome labels), with *softmax* activation |  |  |  |  |
| Number of epochs for training | 25 |  |  |  |  |
| Optimizer | SGD(learning_rate = 0.1, momentum = 0.9, decay = 0.1/25, nesterov = True) |  |  |  |  |
| Loss function | Binary crossentropy |  |  |  |  |

SDG: stochastic descent gradient.

In addition, we monitored the validation loss: when the validation loss would not improve in one epoch, then the learning rate was multiplied by 0.1. We also specified an early stop: when the validation loss would not improve for ten epochs, the training would stop. To choose between the five candidate models we did not implement any data augmentation methods.

We chose the model with the best performance (Table 1 in the main text), which was further tuned (Table 2 in main text).

### Model Architecture

The chosen model (NASNetLarge) presented in this article is a state-of-the-art convolutional neural network (CNN) pre-trained with the ImageNet dataset of images. The NASNetLarge neural network is widely used in computer vision. It is composed of several layers of convolutional cells that extract the most important features from each image, learning which features are the most characteristic from each image category. Most pre-trained state-of-the-art CNNs, such as the AlexNet or the ResNet50, base their innovations in the use of complex activation functions, efficient designs and special layers, such as the Batch Normalization,^4^ which not only improve accuracy performance, but also reduce the time needed to train a model. The ResNet50 neural network, for example, introduces the concept of residual neural networks, layers that skip their connections to other layers by adding connection shortcuts, thus reducing backpropagation training time and better generalizing models.

However, most of these CNNs had complex design that was built by using trial-and-error methods, oblivious to modern state-of-the-art optimization methods such as the use of evolutionary and nature-inspired algorithms or Reinforcement Learning (RL). This is where Neural Search Architecture (NAS) Networks come in play.^5^ Dubbed as a breakthrough in machine learning automation, NAS networks use *AI for AI*. The network’s whole architecture is designed by optimization algorithms, such as Gradient-based search and RL. The idea behind this is that, setting the right restrictions to avoid repetitive inclusion of layers, the network cannot only be trained by its parameters, but by its own architecture, adding and changing layers, activations functions and connections.

Our research uses the NASNetLarge model available by Keras,^1^ pre-trained on the ImageNet dataset on 1000 categories. Because we only have two categories to train on (moderate and extreme), we started by replacing the network’s fully connected classification layer by a 2-neurons layer, with a softmax activation function.^1^ The rest of the original NASNetLarge network was kept unmodified to preserve its proven architecture.

### Model Training

To train our model we used transfer learning. Based on the pre-trained NASNet model, we retained its parameters and froze them, keeping just trainable the last fully connected layer, initializing its parameters randomly by He uniform initialization.^6^ This single-stage training took advantage of the pre-trained parameters speeding up training by just focusing on the last decision layer. The model was trained for 30 epochs. Each epoch is understood as a complete training cycle through the whole train dataset. Data is then fed by batches; once all batches are loaded, an epoch is finished. By using a batch size of 32, training and testing sets are fed to the neural network. The loss function, as we are dealing with a binary classification model, is binary cross entropy.

As optimizer we used the Stochastic Gradient Descent (SGD).^7^ One of the key advantages of this optimizer is due to its stochasticity in selecting each batch for the backpropagation step. Although the model takes longer to converge, unlike other optimizers, the SGD has been proven to converge better local optima, searching for global optima with much more easiness. This factor helps also to reduce overfitting.

### Activation Heatmap by GradCam Visualization

Model interpretability is a feature that has gained importance since the appearance of methods such as GradCam.^8^ This technique uses the values from the gradients in the model’s final feature layer to produce visual explanations, highlighting regions of importance taken by the model to infer a given input. In other words, this technique informs which areas of the input figure were more relevant to make the final classification.

Regions that represent higher gradient values, accounting for most of the last layer activations and network’s decision, are represented by being coloured closer to the red portion of the spectrum. By comparison, regions with the lowest activations, not adding much information to the network’s final decision, appear as areas closer to the blue portion of the spectrum. These gradient values are taken from the network’s last convolutional layer, right before the last pooling layer.

## Notes

### Competing Interest Statement

The authors have declared no competing interest.

